# Plasma Level of ATPase Inhibitory Factor 1 (IF1) and intrinsic capacity in community-dwelling older adults: Prospective data from the MAPT Study

**DOI:** 10.1101/2022.09.02.22279534

**Authors:** Jaqueline Aragoni da Silva, Laurent O. Martinez, Yves Rolland, Souad Najib, Mikaël Croyal, Bertrand Perret, Nabila Jabrane-Ferrat, Hicham El Costa, Sophie Guyonnet, Bruno Vellas, Philipe de Souto Barreto, the MAPT/DSA group

**Affiliations:** Institut du Vieillissement, Gérontopôle de Toulouse, Centre Hospitalo-Universitaire de Toulouse, 37 allées Jules Guesde, 31000 Toulouse, France; Institut des Maladies Métaboliques et Cardiovasculaires, I2MC, Université de Toulouse, Inserm, Université Toulouse III - Paul Sabatier (UPS), UMR1297, Toulouse, France; CERPOP UMR 1295, University of Toulouse III, INSERM, UPS, 31062, Toulouse, France; Nantes Université, CHU Nantes, CNRS, INSERM, l’institut du Thorax, 44000 Nantes, France; Nantes Université, CHU Nantes, CNRS, INSERM, BioCore, US16, SFR Bonamy, F-44000 Nantes, France; CRNH-Ouest Mass Spectrometry Core Facility, 44000 Nantes, France; Toulouse Institute for Infectious and Inflammatory Diseases (Infinity), INSERM-CNRS-University Toulouse III, Toulouse

**Author notes:** Corresponding Authors: Jaqueline Aragoni da Silva, PhD, Gérontopôle de Toulouse, Institut du Vieillissement, Bâtiment B, 37 Allées Jules Guesde, 31000, Toulouse, France, Laurent Martinez, I2MC, 1 Avenue du Pr. Jean Poulhès BP 84225, 31432 Toulouse, Fance. members are listed in the acknowledgements. E-mails : Yves Rolland Souad Najib Mikaël Croyal Bertrand Perret Nabila Jabrane-Ferrat Hicham El Costa Sophie Guyonnet Bruno Vellas Philipe de Souto Barreto.

**Keywords:** ATP synthase, mitochondria, mitochondrial dysfunction, intrinsic capacity, older adults

## Abstract

**Background:** Intrinsic capacity (IC) is a function-related concept that reflects healthy aging. Identifying IC biomarkers is an essential step to slow down functional declines at early stages. ATPase inhibitory factor 1 (IF1) is a multifaceted protein that regulates mitochondrial oxidative phosphorylation (OXPHOS), thereby affecting cellular energy production.

**Objective:** To investigate the association between plasma levels of IF1 and IC changes over four years in community-dwelling older adults.

**Methods:** Community-dwelling older adults aged over 70 years at risk of cognitive decline from the Multidomain Alzheimer Preventive Trial (MAPT Study) were enrolled in this study. A composite IC score was calculated (ranging from 0 to 100; higher is better) over time using data on the following IC domains: locomotion, psychological dimension, cognition, vitality, and sensory ability (vision and hearing; assessed over one year only). Plasma levels of IF1 were assessed one year after the beginning of the study. Mixed-model linear regression adjusted for confounders was performed.

**Results:** A total of 1090 participants had usable IF1 values (mean age 75.3 ± 4.4 years; 64% females). The mean values of plasma IF1 and IC (4 domains) were 565.62 ± 251.92 ng/mL and 74.85 ± 8.43, respectively. Compared to the lowest quartile, low- and high-intermediate IF1 quartiles were cross-sectionally associated with greater composite IC scores of four domains (β_low-intermediate_, 1.33; 95% CI 0.06– 2.60 and β_high-intermediate_, 1.78; 95% CI 0.49–3.06), and the highest quartile associated with a slower decline in composite IC scores of five domains over one year (β_high_ 1.60; 95% CI 0.06– 3.15). The low- and high-intermediate IF1 quartiles were cross-sectionally associated with greater locomotion (β_low-intermediate_, 2.72; 95% CI 0.36–5.08) and vitality scores (β_high-intermediate_, 1.59; 95% CI 0.06–3.12), respectively.

**Conclusion:** This study is the first to report circulating IF1 levels as a mitochondrial-related biomarker associated with IC composite scores in cross-sectional and prospective analyses in community-dwelling older adults. Further research is needed to confirm these findings, in particular, to determine a potential cut-off defining optimal plasma IF1 levels and to unravel the potential mechanisms that can explain these associations.

## INTRODUCTION

A dynamic and life course approach has been outlined by the World Health Organization (WHO) as a new concept of healthy aging. Well-being can be achieved by improving functional ability, which depends on the interplay between environmental factors and intrinsic capacity (IC). IC, which is a central aspect of this framework, is based on a functional-centered rather than a pathology-centered perspective, composed of all physical and mental attributes of an individual^1^. Locomotion, psychological dimensions, cognition, vitality, and sensory abilities have been proposed as the domains representing IC^2^, allowing monitoring of executive functions across the lifespan. Early identification of IC declines can lead to timely interventions, thereby precluding or delaying disability. Currently, little is known about the underlying biological mechanisms of IC, and further knowledge regarding this topic is likely to help identify early IC decline and, therefore, risk stratification of individuals. In this context, mitochondrial function, as a key player in energy balance, may have an important role in the maintenance of optimal IC levels during aging.

Mitochondria are organelles present in eukaryotic cells and they are involved in several key cellular functions, including calcium signaling, cell proliferation and differentiation, control of oxidative stress, and energy production by oxidative phosphorylation (OXPHOS)^3^. Mitochondrial dysfunction is considered to be a hallmark of aging^4^, being deeply involved in the aging process^5^. Indeed, decreased mitochondrial quality and activity have been associated with cellular senescence, chronic inflammation, and a decline in stem cell activity^5^.

ATPase inhibitory factor 1 (IF1) is a nuclear-encoded endogenous inhibitor of the mitochondrial ATP synthase, the fifth complex of the mitochondrial OXPHOS system, the activity of which tightly regulates mitochondrial bioenergetics^6^. For a long time, IF1 has been considered to be a unidirectional inhibitor of ATP synthase, acting only by inhibition of the hydrolase activity (i.e., ATPase activity) of the enzyme during a hypoxic state^6^. However, recent cellular studies support the possibility that IF1 may also partially inhibit the synthetic activity of ATP synthase (i.e., ATP synthesis) during conditions of normal respiration^6^. IF1 has also been reported to contribute to several other cellular processes related to mitochondrial function, such as mitochondrial quality control (mitophagy), redox balance reactions, and the control of cell fate^6^. These recent observations have expanded the role of IF1 in health and disease-related outcomes, ranging from obesity and diabetes to cardiovascular diseases and cancer^6,7^.

In addition to its localization within mitochondria, IF1 is also found in the systemic circulation in humans^8^. In several population studies, circulating IF1 levels were found to be inversely correlated with metabolic syndrome and cardiovascular risk^9,10^. As a biomarker, it has been proposed that increased circulating IF1 may indicate low mobilization in mitochondria and optimal mitochondrial energy production^6^. In light of this, it is hence conceivable that IF1 may play a role in IC. The present study aimed to explore the cross-sectional and prospective associations of plasma IF1 levels with an IC composite score (as well as each IC domain separately) and its change over time in community-dwelling older adults.

## METHODS

### MAPT Study

The Multidomain Alzheimer Preventive Trial (MAPT Study) is a phase III, multicenter, randomized, placebo-controlled trial aimed at investigating the efficacy of isolated supplementation with omega 3 polyunsaturated fatty acids and isolated multidomain intervention (nutritional counseling, physical activity, and cognitive stimulation) or a combination of the two interventions on the improvement of cognitive function in individuals aged 70 years or older, over three years. After the period of intervention, participants were observationally followed for an additional two years^11^.

### Ethical aspects

The study was approved by the French Ethics Committee located in Toulouse (CPP SOOM II) and authorized by the French Health Authority. Written consent was obtained from all participants. The protocol is registered on the clinical trials database (www.clinicaltrials.gov – NCT00672685)^11^.

### Study population

The target population comprised community-dwelling older adults aged 70 years or older at risk of cognitive decline. Inclusion criteria were individuals who met at least one of the following: spontaneous memory complaint; limitation in at least one instrumental activity of daily living (IADL); and a slow walking speed (4-meter usual walking test performance lower than 0.8 m/s). Exclusion criteria were: older adults with dementia or with a Mini-Mental State Examination (MMSE) score lower than 24; limitation in at least one basic activity of daily living (ADL); diseases that could interfere with participation, and those who had taken omega-3 supplements during the last six months. A total of 1680 individuals were enrolled from 13 memory clinics (health centers) in France and Monaco. The period of recruitment was from May 2008 to February 2011 and the follow-up was completed in April 2016^11^.

### Plasma IF1

Plasma IF1 was measured by liquid chromatography-tandem mass spectrometry (LC-MS/MS)^12^. Briefly, IF1 was quantified in 40 µl -aliquots (EDTA plasma) by trypsin proteolysis and subsequent analysis of a prototypic peptide. The intra- and inter-assay variabilities did not exceed 14.2%. Plasma IF1 (ng/mL) was measured in a total of 1097 out of 1680 participants (65.3 %), at the MAPT one-year visit (hereafter called baseline). For the statistical analysis, plasma IF1 outliers (n=7) were computed as those with values above or below four standard deviations from the mean sample and were excluded. Due to the lack of cut-points to classify plasma IF1 values, it was analyzed both as a continuous variable and as distribution quartiles (lowest (Q1), as the reference category).

### Intrinsic capacity

This work includes data collected annually over four years, from the MAPT one-year visit until the visit at the end of the study^11^; therefore, the baseline data corresponds to the MAPT one-year visit (no IC data obtained before IF1 assessment has been used).

The composite IC^1^ score was calculated using the following domains: *Locomotion* assessed by the Short-Physical Performance Battery (SPPB), which is a set of physical performance tests aimed at evaluating lower extremity function in older people. It comprises three tests: balance, gait speed, and chair rise time. Scores range from zero to 12. The higher the score, the better the performance^13^. *Psychological* evaluated by the Geriatric Depression Scale (GDS-15), which is a screening tool for detecting depression in older adults. The short form consists of 15 items, and scores range from zero to 15. Higher scores indicate worse depressive symptoms^14^. *Cognition*. evaluated by the Mini-Mental State Examination, a test of cognitive functions, with scores ranging from zero to 30 (the higher the score, the better the cognition)^15^. *Vitality*, for which the handgrip strength (Jamar^®^ Hydraulic Handheld Dynamometer; Sammons Preston, Bolingbrook, IL, USA -calculated in kilograms) has been considered a reliable indicator^16^. *Sensory*. based on mean scores for visual and hearing evaluation. The vision component was evaluated using the Monoyer Vision Chart; an assessment of distance visual acuity, with scores ranging from 1 to 10. The hearing component was assessed by the Hearing Handicap Inventory – Screening Version; a screening tool to evaluate hearing impairment, with scores ranging from zero to 40^17^.

To allow the comparability of the scales for different measures, rescaling transformations into the same metric were applied. The GDS and Hearing Handicap Inventory scores were multiplied by -1. Each IC domain was rescaled in values ranging from zero to 100, considering their minimum and maximal possible values, respectively. For vitality, the maximal value of the sample across all time points was taken as the highest possible value. Considering this, IC composite scores could range from zero (worst IC possible) to 100 (best IC possible).

Data on the sensory domain were available for only half of the population (individuals randomized for the MAPT multidomain interventions) and at two time-points: baseline (i.e., same time-point as IF1 measurement) and the first year of follow-up. In this sense, two composite IC scores were operationalized in this study. *Main outcome*: IC composed of four domains (without the sensory domain); *Secondary outcome*: IC composed of five domains (including the sensory domain). Additionally, the change in each IC domain was examined separately using the rescaled values of the original assessment tools.

### Confounders

The following variables were taken into account: socio-demographic data (sex, age, and level of education), body mass index (BMI in kg/m^2^; categories were defined according to the World Health Organization classification^18^), moderate-to-vigorous physical activity (MVPA; continuous variable calculated as metabolic equivalent task minutes per week – MET-min/week), and MAPT group allocation.

### Statistical analysis

Descriptive statistics were applied to characterize participants using the MAPT one-year visit, including means and standard deviation, frequencies, and percentages. Normality of the distribution was verified by the Kolmogorov–Smirnov test along with histogram inspection. Comparison variables at baseline according to IF1 quartiles (385.62, 521.66 and 709.42 ng / mL) were evaluated by One-way Analysis of Variance (ANOVA) or the Kruskal–Wallis rank-sum test (depending on the variable distribution) for quantitative variables. Qualitative variables were compared by the Chi-square test.

Linear mixed models were performed by running lme4 packages for R programming language version 4.1.3 software. This is a model that can consider both fixed- and random-effect terms, taking into account nested or hierarchical structures^19^. Models were built separately for each outcome: IC composed of four domains, IC composed of five domains, and for each of the five IC domains. For each model, participants were considered as a random effect. Unadjusted linear mixed models included plasma IF1 levels (either continuous or as quartiles), time (continuous), and the interaction of these two variables (IF1*time) as fixed effects. Then, confounders were added to the adjusted linear mixed models. A sensitivity analysis was conducted restricted to the placebo group. Model fit was checked based on normality of residuals distribution and heteroscedasticity. For all analyses, the significance level was set at p < 0.05.

## RESULTS

### Characterization of the sample

Table 1 lists the characteristics of both the total sample and according to IF1 quartiles. A total of 1090 participants had plasma IF1 information at baseline. Most of these participants were females (64%), with a mean age of 75.3 ± 4.4 years, and 30% had a university degree. The mean plasma IF1 level was 565.62 ± 251.92 ng/mL. Participants in the lowest IF1 quartile had a lower proportion of females, lower MVPA levels, higher BMI and vitality scores compared to the higher quartiles.

**Table 1.** Characteristics of the sample according to quartiles of IF1 at one year (MAPT study)

| Variables | n | Overall<br>n = 1,090 | IF1 quartiles<sup>a</sup> |  |  |  | p-value<sup>b</sup> |
| --- | --- | --- | --- | --- | --- | --- | --- |
|  |  |  | Low<br>n = 272 | Low-intermediate<br>n = 272 | High-intermediate<br>n = 272 | High<br>n = 273 |  |
| **Sex (females)** | 1.09 | 694 (64%) | 151 (55%) | 160 (59%) | 183 (67%) | 200 (73%) | **<0.001** |
| **Age** | 1.09 | 75.30 (4.36) | 75.41 (4.36) | 75.25 (4.56) | 75.24 (4.08) | 75.30 (4.44) | 0.9 |
| **Level of education** | 1.075 |  |  |  |  |  | 0.9 |
| None |  | 50 (4.7%) | 11 (4.1%) | 15 (5.6%) | 13 (4.9%) | 11 (4.1%) |  |
| Primary education |  | 179 (17%) | 41 (15%) | 44 (16%) | 46 (17%) | 48 (18%) |  |
| Secondary education |  | 351 (33%) | 97 (36%) | 88 (33%) | 86 (32%) | 80 (30%) |  |
| High School |  | 168 (16%) | 37 (14%) | 44 (16%) | 47 (18%) | 40 (15%) |  |
| University |  | 327 (30%) | 85 (31%) | 77 (29%) | 75 (28%) | 90 (33%) |  |
| **BMI** | 1.086 | 26.23 (4.07) | 26.94 (4.27) | 26.56 (4.10) | 25.72 (3.45) | 25.71 (4.26) | **<0.001** |
| **MVPA** | 1.085 | 1,148.00 (529.00,<br>1,997.00) | 1,155.00 (525.00,<br>1,936.00) | 1,047.00 (445.50,<br>1,774.00) | 1,191.00 (612.00,<br>2,146.25) | 1,243.00 (570.00,<br>2,208.00) | **0.020** |
| **Locomotion** | 1.065 | 10.67 (1.76) | 10.47 (1.94) | 10.72 (1.54) | 10.91 (1.44) | 10.56 (2.01) | 0.12 |
| **Psychological** | 1.079 | 3.07 (2.61) | 3.20 (2.77) | 2.99 (2.58) | 2.88 (2.44) | 3.22 (2.65) | 0.5 |
| **Cognitive** | 1.084 | 28.06 (1.81) | 27.99 (1.89) | 27.96 (1.86) | 28.22 (1.72) | 28.09 (1.79) | 0.3 |
| **Vitality** | 985 | 26.75 (9.75) | 27.56 (10.18) | 27.35 (9.31) | 26.49 (9.70) | 25.62 (9.72) | **0.046** |
| **Vision** | 444 | 6.96 (2.08) | 7.03 (1.94) | 7.02 (2.00) | 7.00 (2.16) | 6.79 (2.22) | 0.9 |
| **Hearing** | 467 | 6.98 (7.99) | 6.54 (8.08) | 6.80 (8.07) | 7.32 (7.52) | 7.23 (8.35) | 0.7 |
| **IC 4 domains** | 957 | 74.85 (8.43) | 74.17 (9.58) | 75.48 (7.25) | 75.77 (7.81) | 74.02 (8.80) | 0.3 |
| **IC 5 domains** | 397 | 75.69 (7.65) | 75.56 (8.35) | 76.65 (6.90) | 76.52 (7.15) | 74.14 (7.91) | 0.2 |

<sup>a</sup>Mean (SD), Median (Interquartile range), or Frequency (%)<sup>b</sup>Pearson's Chi-squared test; Kruskal-Wallis rank-sum test
BMI: Body mass index; IC: Intrinsic capacity
Quartiles for IF1: 385.62, 521.66 and 709.42 ng / mL
IC 4 domains: Intrinsic capacity composed of locomotion, psychological, cognition, and vitality domain (prospective data over four years, 3997 and 3930 observations on adjusted and unadjusted models, respectively ); IC 5 domains: Intrinsic capacity composed of locomotion, psychological, cognition vitality, and sensory domain (prospective data over one year, 723 and 714 observations on adjusted and unadjusted models, respectively)

### IF1 and composite scores of intrinsic capacity

The findings from mixed-model linear regression for the association between plasma IF1 levels and composite IC scores cross-sectionally and prospectively are presented in Tables 2 and 3, considering IF1 as distribution quartiles or as a continuous variable, respectively. Cross-sectional plasma IF1 was associated with composite scores of IC (Table 2). Specifically, compared to the lowest IF1 quartile (Q1), intermediate IF1 quartiles (Q2 and Q3) were associated with greater composite IC scores of four domains after adjusting for confounders (β_low-intermediate_, 1.33; 95% CI 0.06– 2.60 and β_high-intermediate_, 1.78; 95% CI 0.49–3.06). Longitudinally, the participants in the highest IF1 quartile (Q4) had slower declines in IC scores of five domains over one year (β_high_, 1.60; 95% CI 0.06–3.15). No association was found between plasma IF1 levels as a continuous variable and composite IC scores of four and five domains, neither cross-sectionally nor prospectively (Table 3). Sensitive analysis was performed for the placebo group (IF1 as a continuous variable and IC as a composite score of four domains), and similar results were found (Supplementary material).

**Table 2.** Mixed-model linear regression analysis of the cross-sectional and prospective association of IF1, considered as distribution quartiles, and intrinsic capacity composite score in non-demented, community-dwelling older adults

| *Outcomes* | **Simple Model** |  |  |  |  | **Adjusted model<sup>a</sup>** |  |  |  |  |
| --- | --- | --- | --- | --- | --- | --- | --- | --- | --- | --- |
| | $\beta$<br>*Estimates* | *std. Error* | *CI* | *Statistic* | *p* | $\beta$<br>*Estimates* | *std. Error* | *CI* | *Statistic* | *p-value* |
| **Cross-sectional** |  |  |  |  |  |  |  |  |  |  |
| **IC 4 domains** |  |  |  |  |  |  |  |  |  |  |
| Low-intermediate quartile (Q2) | 1.06 | 0.76 | -0.43 – 2.54 | 1.40 | 0.162 | 1.33 | 0.65 | 0.06 – 2.60 | 2.06 | **0.040** |
| High-intermediate quartile (Q3) | 1.25 | 0.75 | -0.23 – 2.73 | 1.66 | 0.097 | 1.78 | 0.65 | 0.49 – 3.06 | 2.72 | **0.007** |
| High quartile (Q4) | -0.55 | 0.75 | -2.02 – 0.93 | -0.72 | 0.469 | 0.25 | 0.65 | -1.03 – 1.53 | 0.38 | 0.705 |
| **IC 5 domains** |  |  |  |  |  |  |  |  |  |  |
| Low-intermediate quartile (Q2) | 0.30 | 1.04 | -1.74 – 2.33 | 0.28 | 0.776 | 1.08 | 0.91 | -0.72 – 2.87 | 1.18 | 0.238 |
| High-intermediate quartile (Q3) | 0.97 | 1.02 | -1.04 – 2.98 | 0.94 | 0.346 | 1.57 | 0.91 | -0.22 – 3.37 | 1.72 | 0.086 |
| High quartile (Q4) | -1.62 | 1.01 | -3.61 – 0.38 | -1.59 | 0.112 | -1.11 | 0.91 | -2.90 – 0.69 | -1.21 | 0.226 |
| **Prospective<sup>b</sup>** |  |  |  |  |  |  |  |  |  |  |
| **IC 4 domains** |  |  |  |  |  |  |  |  |  |  |
| Low-intermediate quartile (Q2) | -0.24 | 0.15 | -0.53 – 0.06 | -1.59 | 0.113 | -0.26 | 0.15 | -0.55 – 0.04 | -1.72 | 0.086 |
| High-intermediate quartile (Q3) | 0.00 | 0.15 | -0.29 – 0.29 | 0.01 | 0.990 | 0.01 | 0.15 | -0.29 – 0.30 | 0.05 | 0.963 |
| High quartile (Q4) | -0.09 | 0.15 | -0.38 – 0.21 | -0.58 | 0.562 | -0.11 | 0.15 | -0.40 – 0.19 | -0.71 | 0.476 |
| **IC 5 domains** |  |  |  |  |  |  |  |  |  |  |
| Low-intermediate quartile (Q2) | 1.15 | 0.78 | -0.39 – 2.68 | 1.47 | 0.142 | 1.20 | 0.78 | -0.34 – 2.73 | 1.53 | 0.125 |
| High-intermediate quartile (Q3) | -0.03 | 0.79 | -1.57 – 1.51 | -0.04 | 0.971 | 0.28 | 0.79 | -1.27 – 1.82 | 0.35 | 0.725 |
| High quartile (Q4) | 1.48 | 0.78 | -0.06 – 3.02 | 1.89 | 0.059 | 1.60 | 0.79 | 0.06 – 3.15 | 2.04 | **0.042** |

Quartiles for IF1: 385.62, 521.66 and 709.42 ng / mL
a: adjusted for sex, age, level of education, body mass index, physical activity, and MAPT group allocation;
IC 4 domains: Intrinsic capacity composed of locomotion, psychological, cognition, and vitality domain (prospective data over four years, 3997 and 3930 observations on adjusted and unadjusted models, respectively); IC 5 domains: Intrinsic capacity composed of locomotion, psychological, cognition vitality, and sensory domain (prospective data over one year, 723 and 714 observations on adjusted and unadjusted models, respectively);
b: Prospective analyses were based on a four-year follow-up for IC 4 domains and one-year follow-up for IC 5 domains.

**Table 3.** Mixed-model linear regression analysis of the cross-sectional and prospective association of IF1, considered as a continuous variable, and intrinsic capacity composite score in non-demented, community-dwelling older adults

| *Outcomes* | **Simple Model** |  |  |  |  | **Adjusted model<sup>a</sup>** |  |  |  |  |
| --- | --- | --- | --- | --- | --- | --- | --- | --- | --- | --- |
| | $\beta$<br>*Estimates* | *std. Error* | *CI* | *Statistic* | *p* | $\beta$<br>*Estimates* | *std. Error* | *CI* | *Statistic* | *p-value* |
| **Cross-sectional** |  |  |  |  |  |  |  |  |  |  |
| IC 4 domains | -1.53 | 1.06 | -3.62 – 0.55 | -1.44 | 0.149 | -0.56 | 0.93 | -2.40 – 1.27 | -0.6 | 0.547 |
| IC 5 domains | -2.22 | 1.46 | -5.08 – 0.63 | -1.53 | 0.127 | -1.8 | 1.34 | -4.44 – 0.83 | -1.34 | 0.179 |
| **Prospective<sup>b</sup>** |  |  |  |  |  |  |  |  |  |  |
| IC 4 domains | 0.08 | 0.2 | -0.32 – 0.48 | 0.4 | 0.689 | 0.03 | 0.21 | -0.37 – 0.44 | 0.15 | 0.878 |
| IC 5 domains | 1.24 | 1.12 | -0.97 – 3.45 | 1.1 | 0.27 | 1.39 | 1.16 | -0.89 – 3.66 | 1.19 | 0.233 |

<sup>a</sup>: adjusted for sex, age, level of education, body mass index, physical activity, and MAPT group allocation;
IC 4 domains: Intrinsic capacity composed of locomotion, psychological, cognition, and vitality domain (prospective data over four years, 3997 and 3930 observations on adjusted and unadjusted models, respectively ); IC 5 domains: Intrinsic capacity composed of locomotion, psychological, cognition vitality, and sensory domain (prospective data over one year, 723 and 714 observations on adjusted and unadjusted models, respectively);
b: Prospective analyses were based on a four-year follow-up for IC 4 domains and one-year follow-up for IC 5 domains.

### IF1 and each domain of intrinsic capacity (individually)

Tables 4 and 5 present the findings from mixed-model linear regression for the cross-sectional and prospective associations between plasma IF1 levels, considered as distribution quartiles or as a continuous variable, respectively, and each domain of IC individually. Compared to the low IF1 quartile (Q1), the low- and high-intermediate IF1 quartile (Q2 and Q3, respectively) were associated with greater locomotion scores in cross-sectional comparisons (β_low-intermediate_, 3.02; 95% CI 0.44– 5.60 and β_high-intermediate_, 3.29; 95% CI 0.72–5.86). This association remained significant for the low-intermediate IF1 quartile after adjusting for confounders (β_low-intermediate_, 2.72; 95% CI 0.36–5.08). High-intermediate IF1 quartile (Q3), compared to the low IF1 quartile (Q1), was associated with greater vitality scores (β_high-intermediate_, 1.59; 95% CI 0.06– 3.12), after the inclusion of cofounders in the model (Table 4). When analyzed as a continuous variable, plasma IF1 levels were not associated with any of the IC domains in adjusted analysis, neither cross-sectionally nor prospectively (Table 5). However, there was a trend for the prospective association between IF1 and the sensory domain (β, 4.38; 95% CI -0.07–8.82, p = 0.053, Table 5).

**Table 4.** Mixed-model linear regression analysis of the cross-sectional and prospective association of IF1, considered as distribution quartiles, and intrinsic capacity domains in non-demented, community-dwelling older adults

| Outcomes | Simple Model |  |  |  |  | Adjusted model<sup>a</sup> |  |  |  |  |
| --- | --- | --- | --- | --- | --- | --- | --- | --- | --- | --- |
| | $\beta$<br>Estimates | std.<br>Error | CI | Statistic | p | $\beta$<br>Estimates | std. Error | CI | Statistic | p-value |
| **Cross-sectional** |  |  |  |  |  |  |  |  |  |  |
| **Locomotion** |  |  |  |  |  |  |  |  |  |  |
| Low-intermediate quartile (Q2) | 3.02 | 1.32 | 0.44 – 5.60 | 2.3 | **0.022** | 2.72 | 1.20 | 0.36 – 5.08 | 2.26 | **0.024** |
| High-intermediate quartile (Q3) | 3.29 | 1.31 | 0.72 – 5.86 | 2.51 | **0.012** | 2.14 | 1.21 | -0.24 – 4.52 | 1.76 | 0.078 |
| High quartile (Q4) | 0.33 | 1.31 | -2.24 – 2.90 | 0.25 | 0.800 | -0.5 | 1.21 | -2.87 – 1.88 | -0.41 | 0.683 |
| **Psychological** |  |  |  |  |  |  |  |  |  |  |
| Low-intermediate quartile (Q2) | 1.02 | 1.53 | -1.97 – 4.01 | 0.67 | 0.505 | 1.19 | 1.49 | -1.74 – 4.11 | 0.79 | 0.427 |
| High-intermediate quartile (Q3) | 2.12 | 1.52 | -0.87 – 5.11 | 1.39 | 0.164 | 2.46 | 1.51 | -0.51 – 5.42 | 1.62 | 0.104 |
| High quartile (Q4) | -0.11 | 1.52 | -3.10 – 2.87 | -0.07 | 0.940 | -0.27 | 1.51 | -3.23 – 2.69 | -0.18 | 0.858 |
| **Cognition** |  |  |  |  |  |  |  |  |  |  |
| Low-intermediate quartile (Q2) | 0.1 | 0.52 | -0.92 – 1.12 | 0.19 | 0.847 | 0.23 | 0.49 | -0.74 – 1.19 | 0.46 | 0.647 |
| High-intermediate quartile (Q3) | 0.78 | 0.52 | -0.23 – 1.80 | 1.51 | 0.130 | 0.75 | 0.5 | -0.22 – 1.73 | 1.51 | 0.131 |
| High quartile (Q4) | 0.46 | 0.52 | -0.56 – 1.47 | 0.88 | 0.378 | 0.34 | 0.5 | -0.64 – 1.32 | 0.68 | 0.494 |
| **Vitality** |  |  |  |  |  |  |  |  |  |  |
| Low-intermediate quartile (Q2) | 0.05 | 1.17 | -2.25 – 2.34 | 0.04 | 0.968 | 0.91 | 0.77 | -0.60 – 2.43 | 1.18 | 0.237 |
| High-intermediate quartile (Q3) | -0.90 | 1.17 | -3.19 – 1.39 | -0.77 | 0.441 | 1.59 | 0.78 | 0.06 – 3.12 | 2.03 | **0.042** |
| High quartile (Q4) | -2.39 | 1.17 | -4.68 – -0.10 | -2.05 | **0.041** | 1.32 | 0.78 | -0.21 – 2.85 | 1.69 | 0.091 |
| **Sensory** |  |  |  |  |  |  |  |  |  |  |
| Low-intermediate quartile (Q2) | -0.17 | 2.03 | -4.16 – 3.82 | -0.08 | 0.933 | -0.03 | 1.99 | -3.94 – 3.88 | -0.02 | 0.987 |
| High-intermediate quartile (Q3) | -0.57 | 2.00 | -4.49 – 3.35 | -0.29 | 0.775 | -0.98 | 1.99 | -4.88 – 2.92 | -0.49 | 0.622 |
| High quartile (Q4) | -2.32 | 2.00 | -6.24 – 1.61 | -1.16 | 0.247 | -3.22 | 1.99 | -7.13 – 0.70 | -1.61 | 0.107 |
| **Prospective<sup>b</sup>** |  |  |  |  |  |  |  |  |  |  |
| **Locomotion** |  |  |  |  |  |  |  |  |  |  |
| Low-intermediate quartile (Q2) | -0.33 | 0.33 | -0.98 – 0.31 | -1.02 | 0.310 | -0.35 | 0.33 | -1.00 – 0.29 | -1.07 | 0.285 |
| High-intermediate quartile (Q3) | -0.18 | 0.33 | -0.82 – 0.46 | -0.54 | 0.588 | -0.14 | 0.33 | -0.78 – 0.51 | -0.41 | 0.679 |
| High quartile (Q4) | -0.31 | 0.33 | -0.95 – 0.34 | -0.94 | 0.347 | -0.30 | 0.33 | -0.95 – 0.34 | -0.92 | 0.358 |
| **Psychological** |  |  |  |  |  |  |  |  |  |  |
| Low-intermediate quartile (Q2) | -0.42 | 0.33 | -1.06 – 0.22 | -1.28 | 0.200 | -0.41 | 0.33 | -1.05 – 0.23 | -1.25 | 0.212 |
| High-intermediate quartile (Q3) | 0.13 | 0.32 | -0.50 – 0.76 | 0.4 | 0.688 | 0.11 | 0.33 | -0.53 – 0.75 | 0.33 | 0.740 |
| High quartile (Q4) | -0.08 | 0.32 | -0.72 – 0.56 | -0.25 | 0.806 | -0.09 | 0.33 | -0.73 – 0.55 | -0.27 | 0.790 |
| **Cognition** |  |  |  |  |  |  |  |  |  |  |
| Low-intermediate quartile (Q2) | -0.15 | 0.14 | -0.42 – 0.12 | -1.1 | 0.273 | -0.15 | 0.14 | -0.42 – 0.12 | -1.09 | 0.275 |
| High-intermediate quartile (Q3) | -0.14 | 0.14 | -0.41 – 0.12 | -1.04 | 0.296 | -0.12 | 0.14 | -0.39 – 0.15 | -0.90 | 0.370 |
| High quartile (Q4) | 0.01 | 0.14 | -0.26 – 0.28 | 0.07 | 0.941 | 0.03 | 0.14 | -0.24 – 0.30 | 0.20 | 0.838 |
| **Vitality** |  |  |  |  |  |  |  |  |  |  |
| Low-intermediate quartile (Q2) | 0.01 | 0.19 | -0.37 – 0.38 | 0.03 | 0.975 | 0.01 | 0.19 | -0.37 – 0.38 | 0.03 | 0.978 |
| High-intermediate quartile (Q3) | 0.32 | 0.19 | -0.06 – 0.69 | 1.66 | 0.097 | 0.34 | 0.19 | -0.03 – 0.72 | 1.8 | 0.072 |
| High quartile (Q4) | -0.01 | 0.19 | -0.39 – 0.36 | -0.06 | 0.952 | -0.04 | 0.19 | -0.41 – 0.33 | -0.2 | 0.841 |
| **Sensory** |  |  |  |  |  |  |  |  |  |  |
| Low-intermediate quartile (Q2) | -1.09 | 1.54 | -4.11 – 1.94 | -0.71 | 0.48 | -0.95 | 1.53 | -3.95 – 2.04 | -0.62 | 0.532 |
| High-intermediate quartile (Q3) | 1.17 | 1.52 | -1.81 – 4.15 | 0.77 | 0.442 | 1.67 | 1.51 | -1.30 – 4.64 | 1.10 | 0.271 |
| High quartile (Q4) | 1.83 | 1.56 | -1.23 – 4.89 | 1.18 | 0.24 | 2.27 | 1.55 | -0.78 – 5.31 | 1.46 | 0.144 |

Quartiles for IF1: 385.62, 521.66 and 709.42 ng / mL
a: adjusted for sex, age, level of education, body mass index, physical activity, and MAPT group allocation; b: Prospective analyses were based on a four-year follow-up (locomotion, psychological, cognition, and vitality ) and one-year follow-up (sensory)

**Table 5.**
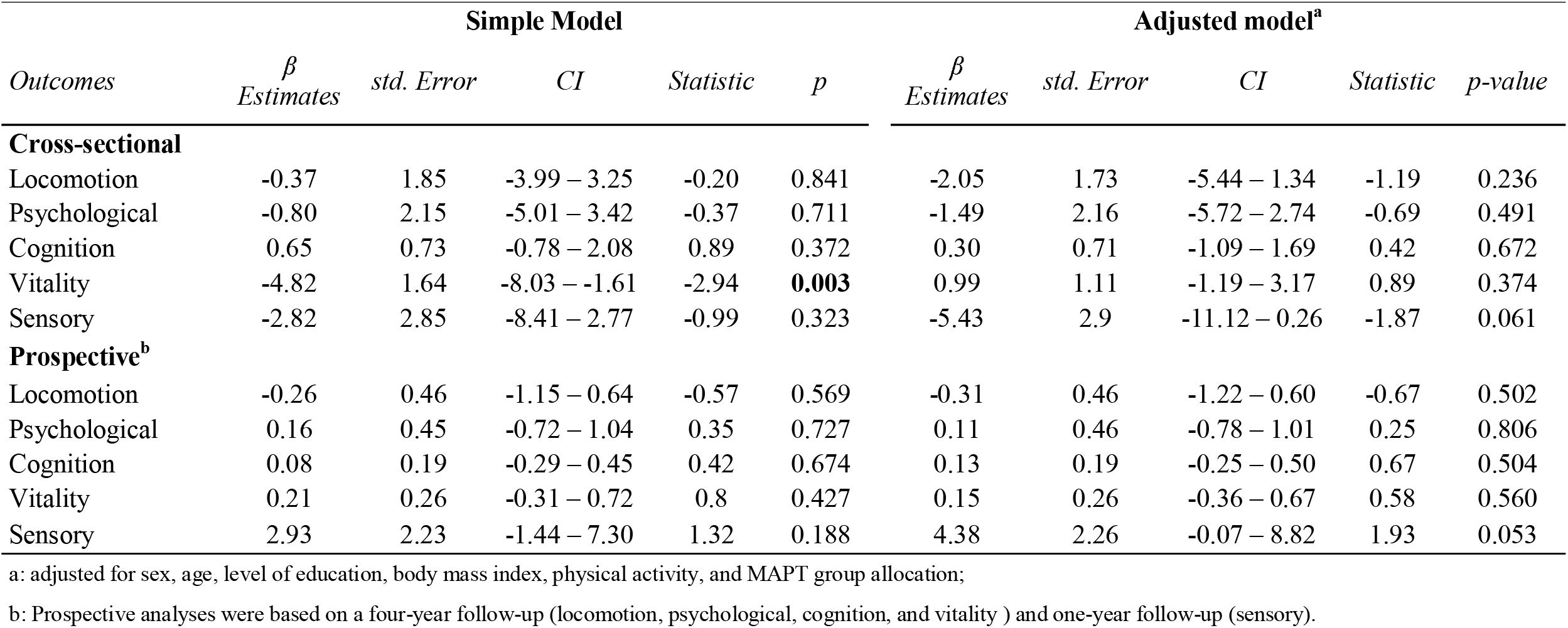
Mixed-model linear regression analysis of the cross-sectional and prospective association of IF1, considered as a continuous variable, and intrinsic capacity domains in non-demented, community-dwelling older adults

| *Outcomes* | **Simple Model** |  |  |  |  | **Adjusted model<sup>a</sup>** |  |  |  |  |
| --- | --- | --- | --- | --- | --- | --- | --- | --- | --- | --- |
| | $\beta$<br>*Estimates* | *std. Error* | *CI* | *Statistic* | *p* | $\beta$<br>*Estimates* | *std. Error* | *CI* | *Statistic* | *p-value* |
| **Cross-sectional** |  |  |  |  |  |  |  |  |  |  |
| Locomotion | -0.37 | 1.85 | -3.99 – 3.25 | -0.20 | 0.841 | -2.05 | 1.73 | -5.44 – 1.34 | -1.19 | 0.236 |
| Psychological | -0.80 | 2.15 | -5.01 – 3.42 | -0.37 | 0.711 | -1.49 | 2.16 | -5.72 – 2.74 | -0.69 | 0.491 |
| Cognition | 0.65 | 0.73 | -0.78 – 2.08 | 0.89 | 0.372 | 0.30 | 0.71 | -1.09 – 1.69 | 0.42 | 0.672 |
| Vitality | -4.82 | 1.64 | -8.03 – -1.61 | -2.94 | **0.003** | 0.99 | 1.11 | -1.19 – 3.17 | 0.89 | 0.374 |
| Sensory | -2.82 | 2.85 | -8.41 – 2.77 | -0.99 | 0.323 | -5.43 | 2.9 | -11.12 – 0.26 | -1.87 | 0.061 |
| **Prospective<sup>b</sup>** |  |  |  |  |  |  |  |  |  |  |
| Locomotion | -0.26 | 0.46 | -1.15 – 0.64 | -0.57 | 0.569 | -0.31 | 0.46 | -1.22 – 0.60 | -0.67 | 0.502 |
| Psychological | 0.16 | 0.45 | -0.72 – 1.04 | 0.35 | 0.727 | 0.11 | 0.46 | -0.78 – 1.01 | 0.25 | 0.806 |
| Cognition | 0.08 | 0.19 | -0.29 – 0.45 | 0.42 | 0.674 | 0.13 | 0.19 | -0.25 – 0.50 | 0.67 | 0.504 |
| Vitality | 0.21 | 0.26 | -0.31 – 0.72 | 0.8 | 0.427 | 0.15 | 0.26 | -0.36 – 0.67 | 0.58 | 0.560 |
| Sensory | 2.93 | 2.23 | -1.44 – 7.30 | 1.32 | 0.188 | 4.38 | 2.26 | -0.07 – 8.82 | 1.93 | 0.053 |

a: adjusted for sex, age, level of education, body mass index, physical activity, and MAPT group allocation;
b: Prospective analyses were based on a four-year follow-up (locomotion, psychological, cognition, and vitality ) and one-year follow-up (sensory).

## DISCUSSION

This study is the first to investigate the association between a mitochondria-related biomarker and IC composite score change over time in older adults. The results show that individuals whose IF1 level was in the highest quartile had slower IC declines compared to those with IF1 levels in the lowest quartile, independently of sex, age, level of education, body mass index, physical activity, or interventional group allocation. In cross-sectional comparisons, intermediate IF1 quartiles (i.e., Q2 and Q3) were cross-sectionally associated with greater IC composite scores.

This study is the first to report circulating IF1 for predicting IC decline. At present, it is only possible to hypothesize regarding the mechanisms linking IF1 and IC decline. Indeed, recent cellular studies support the possibility that IF1 may promote mitochondrial-mediated mechanisms of aging. First, in several human cell types such as skeletal muscle cells, neurons, pancreatic β cells, and hepatocytes, mitochondrial IF1 was reported to reprogram energy metabolism towards enhanced aerobic glycolysis as a result of inhibition of ATP synthase during conditions of normal respiration^20–23^. Accordingly, inhibition of IF1 improves severe OXPHOS dysfunction in human cells^24^. This effect of IF1 on energy metabolism reprogramming is likely to favor the aging process through alterations of metabolic homeostasis, chronic inflammation, and impaired immune responses to environmental and metabolic stresses. Second, it appears that the action of IF1 in restraining OXPHOS activity is associated with the regulation of mitochondria-derived products in a manner that would drive the aging process, by increasing reactive oxygen species (ROS) and by decreasing the NAD+/NADH ratio and α-ketoglutarate (α-KG)^22,25,26^. Thus, there is increasing evidence that mitochondrial IF1 regulates mitochondrial energy metabolism and oxidative stress at a whole-organism level, and may contribute in certain circumstances and organs to mitochondrial dysfunction, and hence contribute to the process of aging. At present, it can be hypothesized that, as a prognostic biomarker of IC decline, circulating IF1 is a surrogate of mitochondrial energetics, with a high level in plasma indicating low mobilization of IF1 in mitochondria and optimal OXPHOS activity^6^.

However, considering the fact that IF1 as quartiles, but not as a continuous variable, was associated with IC, future studies are required to explore optimal cut-off points to determine the threshold at which plasma IF1 would reflect a detrimental situation. Given that only the intermediate IF1 quartiles (Q2 and Q3) but not the highest (Q4) were associated with greater IC composite scores in cross-sectional analysis, it should be considered whether high plasma IF1 level may reflect a reduced mitochondrial resilience in some specific individual conditions. In particular, the IF1 level could have a non-linear or hermetic dose-response relationship with IC decline in older adults. This hypothesis can be connected to a number of emerging concepts and mechanisms of resilience in aging, including mitohormesis, where mild mitochondrial stress leads to metabolic adaptation and improvement of stress defenses that contribute to a delay in age-related diseases and an extended lifespan. Another adaptive mechanism that could be mentioned is the “energy-splicing resilience axis” recently conceptualized and explored by Luigi Ferrucci and colleagues^27^. In this mechanism, a degraded mitochondrial energy status stimulates the production of alternative RNA splicing variants as an adaptive response that preserves energy homeostasis with aging^27^. It could thus be hypothesized that, at some point, high IF1 levels may be associated with reduced mitochondrial resilience.

In addition, the present study indicates that the plasma IF1 level may reflect different domains of IC, such as locomotion, vitality and sensory domains, for which association with IF1 was observed. Low-and high-intermediate plasma IF1 levels were associated with greater locomotion and vitality scores, respectively, in cross-sectional comparisons. However, no longitudinal association was found, and a reverse causal association cannot be ruled out. It has been postulated that IF1 is secreted from skeletal muscle after physical exercise. Exercise-induced circulating IF1 levels have been positively correlated with muscle mass and negatively with age and body fat. It may constitute a myokine that exerts an antidiabetic action, and a potential target to improve physical inactivity-related metabolic diseases^28^. Hence, further studies are needed to explore whether IF1 improves locomotion and handgrip strength or vice versa. Altered IF1 levels have been related to higher apoptosis and neuroinflammation in the brain and retina, playing a role in visual impairment in zebrafish larvae and mice^29^. This is consistent with our results showing that higher plasma IF1 levels are correlated with slower decline in the sensory domain over time.

Furthermore, previous hypotheses have suggested that IF1 should be a potential target for preventing cognitive decline, being highly expressed in neurons, and playing a potential role in neuronal function^30^. For instance, mice with high levels of IF1 exhibit better synaptic transmission and learning, while those with low levels have impaired memory^23^. In addition, it was shown that IF1 administration in a transgenic mouse model of Parkinson’s disease prevents mitochondrial impairment and improves motor functions^31^. These observations, mostly derived from animal models, were not confirmed in the present study. The lack of significant association in this study may be due to the fact that these findings cannot be extended to older adults with cognition impairment. Another possible reason for this discrepancy is that the cognition assessment was not sensitive enough to confirm an association with IF1.

The aforementioned findings support the hypothesis that IF1 may play a role at the whole-organism level and may be involved in the IC decline in older adults. However, the lack of association between IF1 and most of the IC domains individually needs further research to verify whether they share any particularity that is not captured by individual tests. To date, no studies have described the association between IF1 and psychological aspects, hearing, and handgrip strength. This is an emerging topic, and further research will help to clarify the gaps that are still uncovered. As briefly discussed, IF1 does not have a single biological role, and there are still some conflicting results^6^. Most studies to date have been conducted on animals, and little is known about the translation of the findings to humans. In light of this, further investigations are needed to foster discoveries and efforts to define the possible role of IF1 and distinguish between its beneficial and detrimental effects.

### Strength and limitations

This is the first investigation linking the plasma IF1 level to IC, thereby extending the scant literature on this topic. Furthermore, it included a cross-sectional and longitudinal design, allowing investigation of the associations of IF1 with IC change over time. Additionally, state-of-the-art techniques were used in this study, including the liquid chromatography-tandem mass spectrometry analytical method for IF1 assessment^12^, and sensitivity analysis was conducted to assess whether the findings could be different when considering only the placebo group. However, some limitations should also be mentioned. IF1 levels were only available for the MAPT one-year visit and did not include all of the participants from the MAPT study. The lack of prospective data on IF1 precludes investigation of changes in IF1 and its association with changes in IC. No information was available for the sensory domain over four-years follow-up; a composite score of four domains was calculated instead. The sample included individuals aged 70 years or older with at least one condition related to the risk of cognitive decline. Therefore, our results should be interpreted with a degree of caution as they may not apply to the broader population.

## CONCLUSION

IC is a highly valuable assessment to prevent dependence and promote autonomy in older adults. To the best of our knowledge, this is the first study to explore the association between plasma IF1 levels and IC decline. The present study showed that IF1 may be a potential predictor of IC decline over time in non-demented community-dwelling older adults. In particular, intermediate and high IF1 quartiles were cross-sectionally associated with greater IC and with slower decline over one year, respectively. Further investigation of this topic including prospective studies with multiple time-points in more representative samples is needed to confirm whether IF1 is capable of predicting IC declines over the lifespan. Moreover, the mechanisms that may explain these associations still need to be determined. Thus, it will be valuable to elucidate age-related biomarkers that can inform regarding IC impairment as soon as possible and contribute to the development of therapeutic strategies and more optimized interventions to maintain healthy aging and prevent dependency. Studies on this topic are likely to provide relevant contributions to the field.

## Data Availability

All data produced in the present study are available upon reasonable request to the authors

## Acknowledgments

The present work was performed in the context of the Inspire Program, a research platform supported by grants from the Region Occitanie/Pyrénées-Méditerranée (Reference number: 1901175) and the European Regional Development Fund (ERDF) (Project number: MP0022856). Funding was received from Alzheimer Prevention in Occitania and Catalonia (APOC Chair of Excellence - Inspire Program) and Saint Louis University.

The MAPT study was supported by grants from the Gérontopôle of Toulouse, the French Ministry of Health (PHRC 2008, 2009), Pierre Fabre Research Institute (manufacturer of the omega-3 supplement), ExonHit Therapeutics SA, and Avid Radiopharmaceuticals Inc. The promotion of this study was supported by the University Hospital Center of Toulouse. The data sharing activity was supported by the Association Monegasque pour la Recherche sur la maladie d’Alzheimer (AMPA) and the INSERM-University of Toulouse III UMR 1295 (CERPOP) Research Unit.

## MAPT/DSA Group

### MAPT Study Group

Principal investigator: Bruno Vellas (Toulouse); Coordination: Sophie Guyonnet; Project leader: Isabelle Carrié; CRA: Lauréane Brigitte; Investigators: Catherine Faisant, Françoise Lala, Julien Delrieu, Hélène Villars; Psychologists: Emeline Combrouze, Carole Badufle, Audrey Zueras; Methodology, statistical analysis and data management: Sandrine Andrieu, Christelle Cantet, Christophe Morin; Multidomain group: Gabor Abellan Van Kan, Charlotte Dupuy, Yves Rolland (physical and nutritional components), Céline Caillaud, Pierre-Jean Ousset (cognitive component), Françoise Lala (preventive consultation) (Toulouse). The cognitive component was designed in collaboration with Sherry Willis from the University of Seattle, and Sylvie Belleville, Brigitte Gilbert and Francine Fontaine from the University of Montreal.

Co-Investigators in associated centres: Jean-François Dartigues, Isabelle Marcet, Fleur Delva, Alexandra Foubert, Sandrine Cerda (Bordeaux); Marie-Noëlle-Cuffi, Corinne Costes (Castres); Olivier Rouaud, Patrick Manckoundia, Valérie Quipourt, Sophie Marilier, Evelyne Franon (Dijon); Lawrence Bories, Marie-Laure Pader, Marie-France Basset, Bruno Lapoujade, Valérie Faure, Michael Li Yung Tong, Christine Malick-Loiseau, Evelyne Cazaban-Campistron (Foix); Françoise Desclaux, Colette Blatge (Lavaur); Thierry Dantoine, Cécile Laubarie-Mouret, Isabelle Saulnier, Jean-Pierre Clément, Marie-Agnès Picat, Laurence Bernard-Bourzeix, Stéphanie Willebois, Iléana Désormais, Noëlle Cardinaud (Limoges); Marc Bonnefoy, Pierre Livet, Pascale Rebaudet, Claire Gédéon, Catherine Burdet, Flavien Terracol (Lyon), Alain Pesce, Stéphanie Roth, Sylvie Chaillou, Sandrine Louchart (Monaco); Kristel Sudres, Nicolas Lebrun, Nadège Barro-Belaygues (Montauban); Jacques Touchon, Karim Bennys, Audrey Gabelle, Aurélia Romano, Lynda Touati, Cécilia Marelli, Cécile Pays (Montpellier); Philippe Robert, Franck Le Duff, Claire Gervais, Sébastien Gonfrier (Nice); Yannick Gasnier and Serge Bordes, Danièle Begorre, Christian Carpuat, Khaled Khales, Jean-François Lefebvre, Samira Misbah El Idrissi, Pierre Skolil, Jean-Pierre Salles (Tarbes).

MRI group: Carole Dufouil (Bordeaux), Stéphane Lehéricy, Marie Chupin, Jean-François Mangin, Ali Bouhayia (Paris); Michèle Allard (Bordeaux); Frédéric Ricolfi (Dijon); Dominique Dubois (Foix); Marie Paule Bonceour Martel (Limoges); François Cotton (Lyon); Alain Bonafé (Montpellier); Stéphane Chanalet (Nice); Françoise Hugon (Tarbes); Fabrice Bonneville, Christophe Cognard, François Chollet (Toulouse).

PET scans group: Pierre Payoux, Thierry Voisin, Julien Delrieu, Sophie Peiffer, Anne Hitzel, (Toulouse); Michèle Allard (Bordeaux); Michel Zanca (Montpellier); Jacques Monteil (Limoges); Jacques Darcourt (Nice).

Medico-economics group: Laurent Molinier, Hélène Derumeaux, Nadège Costa (Toulouse).

Biological sample collection: Bertrand Perret, Claire Vinel, Sylvie Caspar-Bauguil (Toulouse).

Safety management: Pascale Olivier-Abbal

DSA Group: Sandrine Andrieu, Christelle Cantet, Nicola Coley.

