## Supplementary Material 1 for "Plasma Level of ATPase Inhibitory Factor 1 (IF1) and intrinsic capacity in community-dwelling older adults: Prospective data from the MAPT Study"

| **S1.** Mixed-model linear regression analysis of the cross-sectional and prospective association of IF1, considered as a continuous variable, and intrinsic capacity composite score of 4 domains for the placebo group in non-demented, community-dwelling older adults | | | | | | | | | | | |
| --- | --- | --- | --- | --- | --- | --- | --- | --- | --- | --- | --- |
|  | **Simple Model** | | | | |  | **Adjusted model^a^** | | | | |
| *Outcomes* | *β Estimates* | *std. Error* | *CI* | *Statistic* | *p* |  | *β Estimates* | *std. Error* | *CI* | *Statistic* | *p-value* |
| **Cross-sectional** | 2.08 | 1.84 | -1.53 – 5.69 | 1.13 | 0.258 |  | 2.38 | 1.67 | -0.90 – 5.66 | 1.43 | 0.154 |
| **Prospective^b^** | -0.40 | 0.37 | -1.13 – 0.33 | -1.07 | 0.287 |  | -0.46 | 0.38 | -1.19 – 0.28 | -1.22 | 0.223 |
| ^a^: adjusted for sex, age, level of education, body mass index, and physical activity; | | | | | | | | | | | |
| IC 4 domains: Intrinsic capacity composed of locomotion, psychological, cognition, and vitality domain (prospective data over four years, 1026 and 1012 observations on adjusted and unadjusted models, respectively ). | | | | | | | | | | | |
